# The Cost of Keeping Patients Waiting: Retrospective Treatment-Control Study of Additional Healthcare Utilisation for UK Patients Awaiting Elective Treatment Following COVID-19

**DOI:** 10.1101/2023.07.25.23293143

**Authors:** Charlotte James, Rachel Denholm, Richard Wood

## Abstract

**Objective:** The COVID-19 pandemic has led to increased waiting times for elective treatments in many countries. This study seeks to address a deficit in the literature concerning the effect of long waits on the wider consumption of healthcare resources.

**Methods:** We carried out a retrospective treatment-control study in a healthcare system in South West England from 15 June 2021 to 15 December 2021. We compared weekly contacts with health services of patients waiting over 18 weeks for treatment (‘Treatments’) and people not on a waiting list (‘Controls’). Controls were matched to Treatments based on age, sex, deprivation and multimorbidity. Treatments were stratified by the clinical specialty of the awaited treatment, with healthcare usage assessed over various healthcare settings. T-tests assessed whether there was an increase in healthcare utilisation and bootstrap resampling was used to estimate the magnitude of any differences.

**Results:** A total of 44,616 patients were waiting over 18 weeks (the constitutional target in England) for treatment during the study period. Evidence suggests increases (p < 0.05) in healthcare utilisation for all specialties. Patients in the Cardiothoracic Surgery specialty had the largest increase, requiring 17.9 [4.3, 33.8] additional contacts with secondary care and 17.3 [-1.1, 34.1] additional prescriptions per year.

**Conclusion:** People waiting for treatment consume higher levels of healthcare than comparable individuals not on a waiting list. These findings are relevant for clinicians and managers in better understanding patient need and reducing harm. Results also highlight the possible ‘false economy’ in failing to promptly resolve long elective waits.

**Highlights:**

1. Long waits for elective care can result in additional healthcare needs to manage symptoms up to the point of definitive treatment. While previous studies indicate some association, these mainly consider only a single elective specialty and are limited in the range of healthcare settings covered.
2. The large number of long-wait pathways produced as a consequence of COVID-19 disruption allows for a more holistic analysis, covering the full range of elective treatment specialties and wider healthcare impacts across primary, secondary, mental health, and community care, as well as emergency service calls and prescriptions.
3. Analysis of 44,616 elective care pathways reveals evidence of increases in wider healthcare consumption additional to that expected for similar patients not awaiting elective treatment. This suggests a ‘false economy’ in failing to promptly resolve elective pathways, which should be reflected by healthcare providers in long-term resource allocation decisions.

## Introduction

In the United Kingdom (UK), rising demand for healthcare has led to consistent increases in the size of waiting lists since 2014. As elective procedures were postponed during the pandemic, COVID-19 has accelerated this growth resulting in a substantial backlog of patients awaiting treatment. In March 2022, there were 6.36 million people in the UK waiting for treatment, an increase of 50% compared to March 2020.^1^ The UK government has announced an additional £8 billion over 3 years (2022-2025), allocated to the National Health Service (NHS) to tackle this backlog^.2,3^ For the purpose of decreasing waiting list size, this additional resource could be invested in: alternative methods for prioritising patients waiting for treatment;^4^ pooling patients into larger surgical regions;^5^ increasing capacity via improving surgical schedules, recruitment or use of the private sector.^6,7^

Alongside waiting list size, the median waiting time increased by more than 3 weeks during the COVID-19 pandemic.^1^ In the UK there is an 18-week referral-to-treatment target.^8^ Pre-COVID, this target was being met for 86% of patients, however by March 2022 this had dropped to 62%.^1^ For NHS services to optimise the value of interventions for tackling the waiting list backlog, not only does the resourcing of procedures need to be considered but also the additional resources consumed by people awaiting treatment.

The objective of this study is to quantify the difference in healthcare utilisation of people waiting over 18 weeks for treatment compared to a matched population not on a waiting list. Our results represent a first step in quantifying the hidden costs of keeping people waiting for treatment which is needed for determining care needs, minimising harm and supporting future strategic planning.

## Methods

### Setting, design and population

We conducted a retrospective observational cohort study in Bristol, North Somerset and South Gloucestershire (BNSSG) healthcare system, which serves an approximate one million resident population in South West England. The treatment group consisted of patients registered with a GP practice in BNSSG that were waiting over 18 weeks for treatment at any point in the period 15^th^ June 2021 to 15^th^ December 2021.

### Data

#### Referral to Treatment

Referral to Treatment (RTT) data was obtained from NHS Digital via BNSSG integrated care system (ICS) as a Commissioning Data Set.^9,10^ Since 2007, this data has been submitted by healthcare providers to NHS Digital Secondary Uses Service (SUS) each month. SUS are responsible for collating the data which is used by health care providers and commissioners for service planning and evaluation.^9^

##### The RTT data represents all patients referred for consultant-led elective care to 18 main specialties

General Surgery; Urology; Trauma and Orthopaedic; Ear Nose and Throat; Ophthalmology; Oral Surgery; Neurosurgical; Plastic Surgery; Cardiothoracic Surgery; General Internal Medicine; Gastroenterology; Cardiology; Dermatology; Respiratory Medicine; Neurology; Rheumatology; Elderly Medicine and Gynaecology. Referrals to specialties outside of these 18 are grouped under ‘Other’ and not included in this analysis. The data includes date of referral, specialty, pathway status (open or closed) and date treatment started for each patient pathway.

#### Attributes and Activity

Patient level data was obtained from the BNSSG System Wide Dataset (SWD).^11^ This dataset, which provides linkable data for 98.3% of the 1.07 million population of BNSSG, has been available since 2019. Primary Care data is obtained via a bespoke extract from general practitioners, collated by OneCare (a GP federation). Sourced from EMIS GP administration systems, the extract contains data on GP attendances and prescriptions. SUS contains information on all NHS acute trust outpatient consultations, inpatient admissions, and emergency department attendances, with detailed data on date of attendance, ward specialty and clinical indications (hereafter referred to as secondary care data). The Mental Health Services Data Set (MHSDS), maintained by NHS Digital, contains data covering community mental health consultations and admitted stays in mental health hospitals. Also maintained by NHS Digital, the Community Services Data Set (CSDS) includes intermediate care admissions and patient visits to and from community service teams. Practice Plus Group, the 111 service provider for BNSSG, provides data on 111 calls and the South West Ambulance Service NHS Foundation Trust (SWASFT) provides data on calls made to the Ambulance Service.

##### The BNSSG SWD contains two tables

The attributes table, generated from primary care data, is updated monthly and contains each person’s current demographic, socio-economic and clinical characteristics. The activity table contains information for all discrete patient contacts over the range of healthcare (points of delivery) within BNSSG ICS: Primary Care; Secondary Care; 111; Ambulance (hereafter referred to as 999); Community Services; Mental Health. Information is linked and pseudonymised by a third party, the NHS Commissioning Support Unit. Person Attributes and Activity are linkable through a unique patient identifier; a pseudonymised version of the NHS number.

Study covariates were obtained from the attributes table and included age, gender, socioeconomic status, and presence of chronic conditions. Socioeconomic status was measured using the Index of Multiple Deprivation (IMD). IMD quantifies the relative deprivation of geographical areas in England. In the SWD, the IMD of a person corresponds to the IMD of the area they live in. Presence of chronic conditions was measured using the Quality and Outcomes Framework (QOF) indicators. These indicators include conditions such as obesity, hypertension and diabetes that lead to a person requiring higher levels of healthcare.

#### Processing

For each person on one of the 18 waiting lists during the study period, a pool of controls was obtained from the SWD. Controls were randomly selected from the general population and matched to those on a waiting list, based on gender, 5-year age band, IMD quantile and QOF indicators. Controls were excluded if they were on any waiting list during the study period.

For the treatment group, an individual study period was derived for each patient based on the time of referral and start of treatment (closed pathways). For the treatment group, follow up began at time of referral + 18 weeks, or study start date if already waiting >18 weeks, and censored at date treatment started or end of the study period.

Mean activity per week, stratified by point of delivery, was determined for each participant. For the treatment group, activity directly associated with the referral, namely the GP appointment on the date of referral and the first outpatient appointment, was not included in the analysis.

Patients in the treatment group can have more than one referral to a waiting list and be on waiting lists for multiple specialties. Where a patient had multiple referrals to one specialty, the first referral to a wait list was used, and start of treatment was defined as when all referrals were closed (i.e. the patient was no longer on the waiting list for that specialty). Specialities were analysed separately, as one patient could be on more than one waiting list.

## Statistical analysis

### Is there a difference in healthcare utilisation?

A one-sided t-test was used to determine whether there was a difference in mean activity between the treatment and control groups. Given the many control candidates for each patient on a waiting list (and the desire to avoid reliance on a single selection), repeat bootstrap sampling (1,000 times, with replacement) was used to produce a multitude of treatment-control pairs for each analysis. The t-test was performed to compare the mean weekly activity of the treatment with the control group. For each of the 18 services and 7 points of delivery this yielded 1,000 p-values. A median p-value of less than 0.05 provided strong evidence of a difference in the mean weekly activity of treatments compared to controls. A 95% confidence interval was obtained by taking the 2.5% and 97.5% percentiles of the 1,000 calculated p-values.

### What is the difference in healthcare utilisation?

To quantify the amount of additional healthcare utilisation we estimated the average difference in mean weekly healthcare contacts between the treatment and control group. We used repeat bootstrap sampling (1,000 times, with replacement). During each iteration we found the difference in mean weekly activity of each patient on the waiting list and a matched control. The median and IQR of the distribution of differences was found for all treatment-control pairs, for each point of delivery and each specialty. Repeating this process 1,000 times resulted in distributions of median and IQR values. To summarise the amount of additional healthcare utilisation we reported the means of these distributions.

## Results

### Population characteristics

#### Treatment group

Of the 49,692 patients waiting over 18 weeks for treatment during the study period, 5076 had no matched controls and were excluded from the analysis. The final population consisted of 44,616 treatments across 18 specialties. Treatment characteristics for each speciality are shown in Table 1. The number of patients on each waiting list varies from 29 for Cardiothoracic Surgery to 6,889 for Trauma and Orthopaedic. Most services have an approximately equal number of male and female treatments, however there are some exceptions: Gynaecology and Rheumatology are predominantly female whereas the Urology Service is predominantly male (100%, 71% and 30% female respectively). The mean age of the treatment groups is 53 +/- 5 (mean +/- standard deviation) years for all services apart from Oral Surgery and Gynaecology where patients tend to be younger (31 +/- 21 and 42 +/- 15 years respectively) and Elderly Medicine which has a mean age of 77 +/- 8 years. Dermatology had the lowest percentage of patients who were from a low socio-economic background (IMD <4 = 7.9%), whilst oral surgery had the highest (33.8%). The mean number of QOF conditions is lowest for Oral Surgery (0.51 +/- 0.86) and highest for Elderly Medicine (1.67 +/- 1.34) reflecting the age difference of treatments across specialties. The number of controls per treatment is also reflective of this age difference, with Oral Surgery having the greatest (756.32 +/- 387.53) and Elderly Medicine having the least (200.38 +/- 334.82).

**Table 1.** Treatment description, stratified by specialty. IMD = Index of Multiple Deprivation. QOF = Quality and Outcomes Framework.

| Wait List | N | Female (%) | Age (Mean[SD]) | IMD <4 (%) | QOF (Mean[SD]) | N Controls (Mean[SD]) |
| --- | --- | --- | --- | --- | --- | --- |
| Cardiology | 2450 | 47.8 | 56.03 (18.44) | 21.71 | 1.39 (1.25) | 356.12 (430.01) |
| Cardiothoracic Surgery | 29 | 48.28 | 49.24 (24.2) | 24.14 | 1.0 (0.95) | 427.21 (453.25) |
| Dermatology Service | 1848 | 58.44 | 49.12 (22.38) | 17.91 | 0.83 (1.06) | 593.53 (437.5) |
| Ear Nose and Throat | 3337 | 58.02 | 49.99 (19.94) | 24.36 | 0.84 (1.04) | 579.81 (438.18) |
| Elderly Medicine | 97 | 49.48 | 77.0 (7.69) | 18.56 | 1.67 (1.34) | 200.38 (334.82) |
| Gastroenterology | 2185 | 50.98 | 53.23 (17.38) | 24.58 | 1.06 (1.12) | 494.29 (442.42) |
| General Internal Medicine | 350 | 53.14 | 49.9 (18.9) | 21.71 | 1.19 (1.18) | 428.0 (447.82) |
| General Surgery | 4335 | 61.96 | 53.51 (17.29) | 22.33 | 1.04 (1.17) | 516.13 (447.12) |
| Gynaecology | 4375 | 99.5 | 41.56 (15.07) | 25.69 | 0.67 (0.87) | 694.41 (400.39) |
| Neurology | 1780 | 59.49 | 46.48 (18.2) | 27.36 | 0.99 (1.05) | 507.24 (447.25) |
| Neurosurgical | 555 | 56.22 | 56.45 (16.07) | 24.14 | 1.09 (1.13) | 476.44 (437.39) |
| Ophthalmology | 5718 | 58.17 | 65.15 (18.71) | 21.3 | 1.28 (1.23) | 370.32 (419.9) |
| Oral Surgery | 4200 | 55.93 | 31.01 (20.74) | 33.83 | 0.51 (0.86) | 756.32 (387.53) |
| Plastic Surgery | 930 | 56.77 | 51.99 (20.9) | 21.18 | 0.84 (1.02) | 563.87 (442.09) |
| Respiratory Medicine | 1342 | 47.17 | 54.22 (17.18) | 26.23 | 1.46 (1.18) | 321.3 (404.62) |
| Rheumatology | 1018 | 71.12 | 49.71 (17.63) | 30.26 | 0.99 (1.05) | 513.62 (442.83) |
| Trauma and Orthopaedic | 6889 | 53.94 | 56.16 (18.0) | 22.44 | 1.08 (1.14) | 477.08 (440.81) |
| Urology | 3178 | 29.61 | 58.11 (18.79) | 22.34 | 1.14 (1.18) | 446.07 (436.18) |

#### Healthcare utilisation

Across the 7 points of delivery, healthcare utilisation in the treatment group is greatest for primary care, prescriptions, and secondary care (Table S1). Patients waiting for Cardiothoracic Surgery have the greatest utilisation with a median of 3 [1, 7.5] contacts with primary care, 15 [3, 33.5] prescriptions and 5.5 [2.25, 9] contacts with secondary care. For community services, mental health, 111 and 999 the overall number of contacts during a treatment’s individual study period were very low: the median number of contacts is 0 for all four specialties (Table S1). Controls had lower healthcare utilisation during the study periods, with prescriptions being the only point of delivery where the median number of contacts is non-zero (Table S2).

### Is there a difference in healthcare utilisation?

There is evidence of a difference in the healthcare utilisation of patients waiting over 18 weeks for treatment compared to matched controls (Table S3). For all specialties patients waiting for treatment show increased utilisation of secondary care (p <= 0.002). Elderly Medicine is the only specialty for which there is very little evidence of an increased use of primary care (p = 0.66 [0.55, 0.67]) and prescriptions (p = 0.32 [0.14,0.61]) across the two groups. There is strong evidence that patients waiting for the General Surgery, Urology, Trauma and Orthopaedic, Neurology and Gynaecology services show increased utilisation across all points of delivery (p<0.01), compared to controls.

### What is the difference in healthcare utilisation?

Patients waiting over 18 weeks for treatment demonstrate higher levels of healthcare utilisation than patients not waiting for treatment (Figure 1, Table S4). The additional healthcare utilisation is greatest for primary care prescriptions, followed by secondary care. Patients waiting for the Cardiothoracic Surgery service have a median of 17.9 [4.3, 33.8] additional contacts with secondary care and 17.3 [-1.1, 34.1] additional prescriptions per year compared to matched controls not waiting for treatment. Patients waiting for this service also show the largest median increase in the number of GP appointments (‘primary care contact’) (5.5 [-0.3, 15.9]), however in this point of delivery at least 25% of people waiting for Cardiothoracic Surgery show no increase in utilisation (25^th^ percentile < 0). Of the 18 specialties, patients waiting for the Oral Surgery, Ophthalmology, Gynaecology and Dermatology services had median increases of 0.0 across all points of delivery: at least 50% of patients waiting for these services had no additional healthcare utilisation in all points of delivery. The 111, 999, community and mental health services show the smallest increases in healthcare utilisation, with the median number of additional contacts per year being 0.0 [0.0,0.0] for all specialties. This suggests that, for these services, where there is evidence of increased utilisation (p<0.01, Table S3, Figure 1) this increase can be attributed to less than 25% of the people waiting for treatment (75^th^ percentile = 0.0).

**Figure 1.**
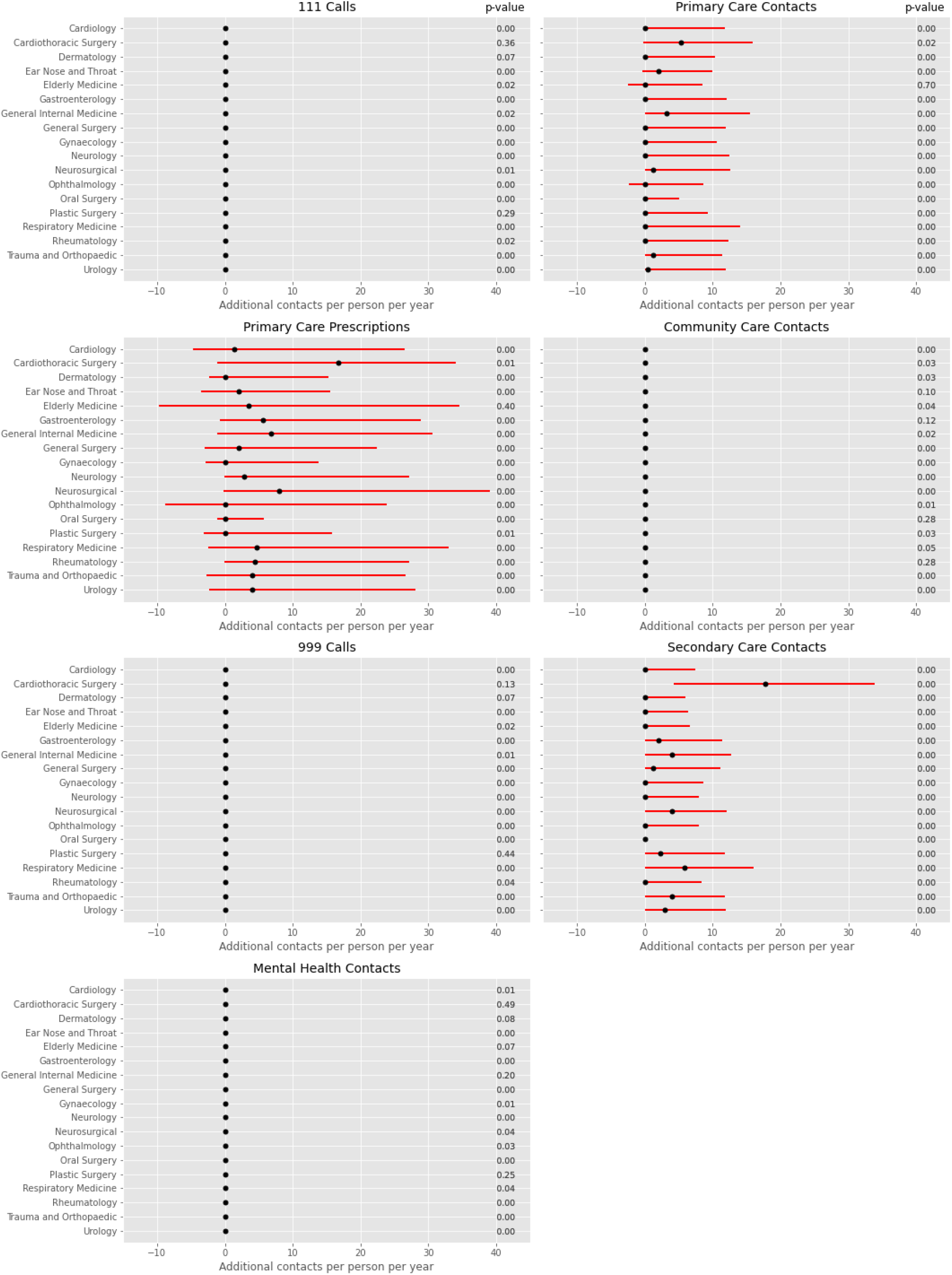
Additional yearly contacts with the health service for patients waiting for treatment (treatments) compared to matched controls. Black dots represent median additional contacts per person per year. Red lines represent the interquartile range. p-values represent evidence for treatments having an increase in yearly contacts. p<0.05 constitutes strong evidence of an increase in healthcare utilisation while waiting for treatment.

## Discussion

Our study marks an initial step in quantifying the hidden cost of increases in waiting list size and waiting times. Using a retrospective cohort study design we have demonstrated that the healthcare utilisation of patients waiting for treatment is greater than a matched population not waiting for treatment. We found evidence of increases in healthcare utilisation across all specialties and all points of delivery. There was strong evidence that there was an increase in the utilisation of secondary care (p<=0.002) for all specialties in the treatment group, compared to controls. The amount of additional utilisation was greatest for primary care prescriptions and secondary care. Patients waiting for the Cardiothoracic Surgery service had the greatest additional utilisation in 3 out of the 7 points of delivery: primary care (5.5 [-0.3, 15.9] additional contacts per year); secondary care (17.9 [4.3, 33.8] additional contacts per year) and prescriptions (17.3 [-1.1, 34.1] additional per year). At least 50% of patients waiting for the Oral Surgery, Ophthalmology, Gynaecology and Dermatology services had no additional healthcare utilisation. Our results demonstrate the burden of increasing waiting times on both patients and health services.

This is one of the first studies to consider the wider implications of long waiting times for patients and health services. From a value-based perspective, knowledge of the amount of extra resource being spent on patients while waiting for treatment is crucial to optimising the cost-effectiveness of any intervention to reduce waiting list size. The objective of our study was to determine whether there was an increase in healthcare utilisation when waiting for treatment. By quantifying the magnitude of this increase, our results will assist strategic planners in assigning a cost to waiting times and waiting list size.

The additional service utilisation of patients waiting over 18 weeks for treatment should be considered as an example of failure-demand within the health service: if patients received treatment earlier, they would not be requiring additional support over a prolonged period. Previous work has demonstrated how failure-demand generated by one component of a health system results in demand being deflected to other components.^12,13^ The consequence is an increase in pressure elsewhere in the system and a negative impact on patient experience.^12^ In combination with our results, this highlights the need for a whole-system approach to tackling the waiting list backlog: a thorough evaluation of interventions, such as increasing surgical capacity and revising existing methods for prioritising patients, requires consideration of the impact on the health service as a whole.

## Conclusion

We have provided evidence that patients waiting for treatment in the UK have higher levels of healthcare utilisation than people not waiting for treatment. Our results can be used to evaluate the cost-effectiveness of interventions to reduce the waiting list backlog. The evidence of increased healthcare utilisation highlights the need for a whole-system approach to tackling the waiting list backlog.

## Data Availability

Data used in this study is not available

## Author Contributions

*Concept and design*: James, Denholm, Wood

*Acquisition of data*: James

*Analysis and interpretation of data*: James

D*rafting of the manuscript*: James

*Critical revision of the paper for important intellectual content*: Denholm, Wood

*Obtaining funding*: Denholm, Wood

*Supervision*: Denholm, Wood

