## Supplement for "The Cost of Keeping Patients Waiting: Retrospective Treatment-Control Study of Additional Healthcare Utilisation for UK Patients Awaiting Elective Treatment Following COVID-19"

### *Health Service Utilisation*

#### Treatments

| | Community | | | |
|---|---|---|---|---|
| **Wait List** | **Mean** | **Median** | **Std** | **IQR** |
| Cardiology Service | 0.63 | 0 | 5.62 | [0.0,0.0] |
| Cardiothoracic Surgery Service | 1.33 | 0 | 3.55 | [0.0,0.0] |
| Dermatology Service | 0.46 | 0 | 4.73 | [0.0,0.0] |
| Ear Nose and Throat Service | 0.50 | 0 | 5.63 | [0.0,0.0] |
| Elderly Medicine Service | 3.06 | 0 | 10.12 | [0.0,1.0] |
| Gastroenterology Service | 0.49 | 0 | 3.48 | [0.0,0.0] |
| General Internal Medicine Service | 0.99 | 0 | 4.53 | [0.0,0.0] |
| General Surgery Service | 1.19 | 0 | 7.84 | [0.0,0.0] |
| Gynaecology Service | 0.34 | 0 | 2.88 | [0.0,0.0] |
| Neurology Service | 0.90 | 0 | 7.04 | [0.0,0.0] |
| Neurosurgical Service | 1.54 | 0 | 7.60 | [0.0,0.0] |
| Ophthalmology Service | 1.02 | 0 | 9.18 | [0.0,0.0] |
| Oral Surgery Service | 0.24 | 0 | 2.72 | [0.0,0.0] |
| Plastic Surgery Service | 0.73 | 0 | 8.16 | [0.0,0.0] |
| Respiratory Medicine Service | 0.65 | 0 | 4.25 | [0.0,0.0] |
| Rheumatology Service | 0.49 | 0 | 4.85 | [0.0,0.0] |
| Trauma and Orthopaedic Service | 0.86 | 0 | 7.51 | [0.0,0.0] |
| Urology Service | 1.25 | 0 | 9.32 | [0.0,0.0] |
| | **Mental Health** | | | |
| | **Mean** | **Median** | **Std** | **IQR** |
| Cardiology Service | 0.17 | 0 | 1.87 | [0.0,0.0] |
| Cardiothoracic Surgery Service | 0.13 | 0 | 0.72 | [0.0,0.0] |
| Dermatology Service | 0.12 | 0 | 0.93 | [0.0,0.0] |
| Ear Nose and Throat Service | 0.24 | 0 | 1.76 | [0.0,0.0] |
| Elderly Medicine Service | 0.38 | 0 | 2.59 | [0.0,0.0] |
| Gastroenterology Service | 0.23 | 0 | 2.36 | [0.0,0.0] |
| General Internal Medicine Service | 0.26 | 0 | 2.24 | [0.0,0.0] |
| General Surgery Service | 0.20 | 0 | 1.75 | [0.0,0.0] |
| Gynaecology Service | 0.20 | 0 | 1.34 | [0.0,0.0] |
| Neurology Service | 0.50 | 0 | 3.13 | [0.0,0.0] |
| Neurosurgical Service | 0.22 | 0 | 1.44 | [0.0,0.0] |
| Ophthalmology Service | 0.13 | 0 | 1.35 | [0.0,0.0] |
| Oral Surgery Service | 0.21 | 0 | 1.80 | [0.0,0.0] |
| Plastic Surgery Service | 0.12 | 0 | 1.02 | [0.0,0.0] |
| Respiratory Medicine Service | 0.20 | 0 | 1.68 | [0.0,0.0] |
| Rheumatology Service | 0.28 | 0 | 1.98 | [0.0,0.0] |
| Trauma and Orthopaedic Service | 0.16 | 0 | 1.50 | [0.0,0.0] |

| | | | | |
|---|---|---|---|---|
| Urology Service | 0.19 | 0 | 1.71 | [0.0,0.0] |
| | 999 | | | |
| | **Mean** | **Median** | **Std** | **IQR** |
| Cardiology Service | 0.11 | 0 | 0.52 | [0.0,0.0] |
| Cardiothoracic Surgery Service | 0.23 | 0 | 0.56 | [0.0,0.0] |
| Dermatology Service | 0.04 | 0 | 0.27 | [0.0,0.0] |
| Ear Nose and Throat Service | 0.08 | 0 | 0.49 | [0.0,0.0] |
| Elderly Medicine Service | 0.22 | 0 | 0.71 | [0.0,0.0] |
| Gastroenterology Service | 0.19 | 0 | 2.18 | [0.0,0.0] |
| General Internal Medicine Service | 0.11 | 0 | 0.43 | [0.0,0.0] |
| General Surgery Service | 0.10 | 0 | 0.50 | [0.0,0.0] |
| Gynaecology Service | 0.07 | 0 | 0.58 | [0.0,0.0] |
| Neurology Service | 0.22 | 0 | 1.74 | [0.0,0.0] |
| Neurosurgical Service | 0.15 | 0 | 0.69 | [0.0,0.0] |
| Ophthalmology Service | 0.08 | 0 | 0.38 | [0.0,0.0] |
| Oral Surgery Service | 0.04 | 0 | 0.26 | [0.0,0.0] |
| Plastic Surgery Service | 0.05 | 0 | 0.29 | [0.0,0.0] |
| Respiratory Medicine Service | 0.11 | 0 | 0.44 | [0.0,0.0] |
| Rheumatology Service | 0.07 | 0 | 0.37 | [0.0,0.0] |
| Trauma and Orthopaedic Service | 0.08 | 0 | 0.45 | [0.0,0.0] |
| Urology Service | 0.12 | 0 | 0.76 | [0.0,0.0] |
| | **111** | | | |
| | **Mean** | **Median** | **Std** | **IQR** |
| Cardiology Service | 0.09 | 0 | 0.45 | [0.0,0.0] |
| Cardiothoracic Surgery Service | 0.07 | 0 | 0.36 | [0.0,0.0] |
| Dermatology Service | 0.05 | 0 | 0.26 | [0.0,0.0] |
| Ear Nose and Throat Service | 0.09 | 0 | 0.41 | [0.0,0.0] |
| Elderly Medicine Service | 0.12 | 0 | 0.45 | [0.0,0.0] |
| Gastroenterology Service | 0.11 | 0 | 0.68 | [0.0,0.0] |
| General Internal Medicine Service | 0.12 | 0 | 0.56 | [0.0,0.0] |
| General Surgery Service | 0.09 | 0 | 0.47 | [0.0,0.0] |
| Gynaecology Service | 0.12 | 0 | 0.79 | [0.0,0.0] |
| Neurology Service | 0.15 | 0 | 0.74 | [0.0,0.0] |
| Neurosurgical Service | 0.11 | 0 | 0.49 | [0.0,0.0] |
| Ophthalmology Service | 0.07 | 0 | 0.35 | [0.0,0.0] |
| Oral Surgery Service | 0.08 | 0 | 0.38 | [0.0,0.0] |
| Plastic Surgery Service | 0.07 | 0 | 0.36 | [0.0,0.0] |
| Respiratory Medicine Service | 0.09 | 0 | 0.40 | [0.0,0.0] |
| Rheumatology Service | 0.07 | 0 | 0.32 | [0.0,0.0] |
| Trauma and Orthopaedic Service | 0.09 | 0 | 0.54 | [0.0,0.0] |
| Urology Service | 0.12 | 0 | 0.60 | [0.0,0.0] |
| | **Primary Care Contact** | | | |
| | **Mean** | **Median** | **Std** | **IQR** |
| Cardiology Service | 3.33 | 1 | 5.10 | [0.0,5.0] |

| | | | | |
|---|---|---|---|---|
| Cardiothoracic Surgery Service | 5.90 | 3 | 7.17 | [1.0,7.5] |
| Dermatology Service | 2.47 | 1 | 4.20 | [0.0,3.0] |
| Ear Nose and Throat Service | 3.84 | 2 | 5.06 | [0.0,5.0] |
| Elderly Medicine Service | 3.53 | 2 | 5.07 | [0.0,5.0] |
| Gastroenterology Service | 3.56 | 2 | 5.27 | [0.0,5.0] |
| General Internal Medicine Service | 4.24 | 2 | 6.16 | [0.0,6.0] |
| General Surgery Service | 4.05 | 2 | 6.58 | [0.0,5.0] |
| Gynaecology Service | 3.09 | 1 | 4.69 | [0.0,4.0] |
| Neurology Service | 3.24 | 1 | 5.25 | [0.0,4.0] |
| Neurosurgical Service | 4.12 | 2 | 5.59 | [0.0,6.0] |
| Ophthalmology Service | 3.20 | 1 | 5.13 | [0.0,4.0] |
| Oral Surgery Service | 1.81 | 0 | 3.44 | [0.0,2.0] |
| Plastic Surgery Service | 2.96 | 1 | 5.74 | [0.0,4.0] |
| Respiratory Medicine Service | 3.99 | 2 | 5.48 | [0.0,6.0] |
| Rheumatology Service | 3.22 | 1 | 4.48 | [0.0,4.0] |
| Trauma and Orthopaedic Service | 4.14 | 2 | 6.04 | [0.0,6.0] |
| Urology Service | 4.01 | 2 | 5.58 | [0.0,6.0] |
| | **Prescription** | | | |
| | **Mean** | **Median** | **Std** | **IQR** |
| Cardiology Service | 14.51 | 5 | 32.10 | [1.0,16.0] |
| Cardiothoracic Surgery Service | 18.13 | 15 | 15.47 | [3.0,33.5] |
| Dermatology Service | 6.99 | 2 | 16.98 | [0.0,7.0] |
| Ear Nose and Throat Service | 12.30 | 4 | 29.38 | [1.0,13.0] |
| Elderly Medicine Service | 19.56 | 12 | 29.55 | [2.0,26.0] |
| Gastroenterology Service | 14.11 | 5 | 32.00 | [1.0,15.0] |
| General Internal Medicine Service | 17.67 | 7 | 39.48 | [1.0,17.0] |
| General Surgery Service | 14.17 | 4 | 34.56 | [1.0,14.0] |
| Gynaecology Service | 6.67 | 2 | 16.92 | [0.0,6.0] |
| Neurology Service | 11.81 | 3 | 32.25 | [0.0,11.0] |
| Neurosurgical Service | 19.44 | 7 | 43.53 | [1.0,19.0] |
| Ophthalmology Service | 15.07 | 6 | 32.11 | [1.0,16.0] |
| Oral Surgery Service | 4.77 | 1 | 16.02 | [0.0,3.0] |
| Plastic Surgery Service | 9.00 | 2 | 20.21 | [0.0,9.0] |
| Respiratory Medicine Service | 16.56 | 7 | 31.47 | [1.0,20.0] |
| Rheumatology Service | 11.70 | 4 | 25.73 | [0.0,12.0] |
| Trauma and Orthopaedic Service | 16.14 | 6 | 34.26 | [1.0,19.0] |
| Urology Service | 16.90 | 6 | 35.34 | [1.0,19.0] |
| | **Secondary** | | | |
| | **Mean** | **Median** | **Std** | **IQR** |
| Cardiology Service | 1.99 | 1 | 5.21 | [0.0,2.0] |
| Cardiothoracic Surgery Service | 8.07 | 5.5 | 10.54 | [2.25,9.0] |
| Dermatology Service | 1.25 | 0 | 2.88 | [0.0,1.0] |
| Ear Nose and Throat Service | 2.13 | 1 | 4.01 | [0.0,3.0] |
| Elderly Medicine Service | 2.18 | 1 | 3.41 | [0.0,3.0] |

| | | | | |
|---|---|---|---|---|
| Gastroenterology Service | 2.58 | 1 | 5.34 | [0.0,3.0] |
| General Internal Medicine Service | 3.22 | 2 | 6.19 | [0.0,4.0] |
| General Surgery Service | 2.94 | 1 | 6.61 | [0.0,3.0] |
| Gynaecology Service | 2.06 | 1 | 3.96 | [0.0,3.0] |
| Neurology Service | 1.96 | 0 | 4.27 | [0.0,2.0] |
| Neurosurgical Service | 2.93 | 1 | 4.49 | [0.0,4.0] |
| Ophthalmology Service | 2.02 | 1 | 3.98 | [0.0,3.0] |
| Oral Surgery Service | 0.78 | 0 | 2.20 | [0.0,1.0] |
| Plastic Surgery Service | 2.27 | 1 | 4.43 | [0.0,3.0] |
| Respiratory Medicine Service | 2.95 | 2 | 4.73 | [0.0,4.0] |
| Rheumatology Service | 1.94 | 0 | 3.69 | [0.0,2.0] |
| Trauma and Orthopaedic Service | 2.75 | 1 | 4.36 | [0.0,4.0] |
| Urology Service | 2.76 | 1 | 4.93 | [0.0,3.75] |

**Table S1** Number of contacts patients waiting over 18 weeks for treatment had with the health service during their individual study periods.

**Controls**

| | Community | | | |
|---|---|---|---|---|
| **Wait List** | **Mean** | **Median** | **Std** | **IQR** |
| Cardiology Service | 0.06 | 0 | 0.87 | [0.0,0.0] |
| Cardiothoracic Surgery Service | 0.19 | 0 | 1.29 | [0.0,0.0] |
| Dermatology Service | 0.05 | 0 | 0.71 | [0.0,0.0] |
| Ear Nose and Throat Service | 0.09 | 0 | 1.11 | [0.0,0.0] |
| Elderly Medicine Service | 0.11 | 0 | 1.69 | [0.0,0.0] |
| Gastroenterology Service | 0.06 | 0 | 1.12 | [0.0,0.0] |
| General Internal Medicine Service | 0.06 | 0 | 0.93 | [0.0,0.0] |
| General Surgery Service | 0.07 | 0 | 1.09 | [0.0,0.0] |
| Gynaecology Service | 0.08 | 0 | 0.84 | [0.0,0.0] |
| Neurology Service | 0.05 | 0 | 0.69 | [0.0,0.0] |
| Neurosurgical Service | 0.07 | 0 | 1.08 | [0.0,0.0] |
| Ophthalmology Service | 0.10 | 0 | 1.60 | [0.0,0.0] |
| Oral Surgery Service | 0.10 | 0 | 0.74 | [0.0,0.0] |
| Plastic Surgery Service | 0.07 | 0 | 1.06 | [0.0,0.0] |
| Respiratory Medicine Service | 0.06 | 0 | 1.01 | [0.0,0.0] |
| Rheumatology Service | 0.06 | 0 | 0.93 | [0.0,0.0] |
| Trauma and Orthopaedic Service | 0.07 | 0 | 1.26 | [0.0,0.0] |
| Urology Service | 0.07 | 0 | 1.28 | [0.0,0.0] |
| | **Mental Health** | | | |
| | **Mean** | **Median** | **Std** | **IQR** |
| Cardiology Service | 0.06 | 0 | 0.82 | [0.0,0.0] |
| Cardiothoracic Surgery Service | 0.11 | 0 | 1.31 | [0.0,0.0] |
| Dermatology Service | 0.07 | 0 | 0.83 | [0.0,0.0] |
| Ear Nose and Throat Service | 0.09 | 0 | 1.02 | [0.0,0.0] |
| Elderly Medicine Service | 0.02 | 0 | 0.44 | [0.0,0.0] |
| Gastroenterology Service | 0.06 | 0 | 0.80 | [0.0,0.0] |
| General Internal Medicine Service | 0.06 | 0 | 0.79 | [0.0,0.0] |
| General Surgery Service | 0.06 | 0 | 0.80 | [0.0,0.0] |
| Gynaecology Service | 0.11 | 0 | 1.09 | [0.0,0.0] |
| Neurology Service | 0.08 | 0 | 0.92 | [0.0,0.0] |
| Neurosurgical Service | 0.05 | 0 | 0.77 | [0.0,0.0] |
| Ophthalmology Service | 0.04 | 0 | 0.68 | [0.0,0.0] |
| Oral Surgery Service | 0.08 | 0 | 0.98 | [0.0,0.0] |
| Plastic Surgery Service | 0.06 | 0 | 0.87 | [0.0,0.0] |
| Respiratory Medicine Service | 0.05 | 0 | 0.72 | [0.0,0.0] |
| Rheumatology Service | 0.08 | 0 | 0.91 | [0.0,0.0] |
| Trauma and Orthopaedic Service | 0.06 | 0 | 0.85 | [0.0,0.0] |
| Urology Service | 0.05 | 0 | 0.76 | [0.0,0.0] |
| | **999** | | | |
| | **Mean** | **Median** | **Std** | **IQR** |
| Cardiology Service | 0.01 | 0 | 0.15 | [0.0,0.0] |

| | | | | |
|---|---|---|---|---|
| Cardiothoracic Surgery Service | 0.03 | 0 | 0.21 | [0.0,0.0] |
| Dermatology Service | 0.01 | 0 | 0.14 | [0.0,0.0] |
| Ear Nose and Throat Service | 0.02 | 0 | 0.18 | [0.0,0.0] |
| Elderly Medicine Service | 0.02 | 0 | 0.14 | [0.0,0.0] |
| Gastroenterology Service | 0.01 | 0 | 0.16 | [0.0,0.0] |
| General Internal Medicine Service | 0.01 | 0 | 0.16 | [0.0,0.0] |
| General Surgery Service | 0.01 | 0 | 0.16 | [0.0,0.0] |
| Gynaecology Service | 0.02 | 0 | 0.19 | [0.0,0.0] |
| Neurology Service | 0.01 | 0 | 0.16 | [0.0,0.0] |
| Neurosurgical Service | 0.01 | 0 | 0.16 | [0.0,0.0] |
| Ophthalmology Service | 0.01 | 0 | 0.16 | [0.0,0.0] |
| Oral Surgery Service | 0.02 | 0 | 0.17 | [0.0,0.0] |
| Plastic Surgery Service | 0.01 | 0 | 0.16 | [0.0,0.0] |
| Respiratory Medicine Service | 0.01 | 0 | 0.16 | [0.0,0.0] |
| Rheumatology Service | 0.01 | 0 | 0.16 | [0.0,0.0] |
| Trauma and Orthopaedic Service | 0.02 | 0 | 0.17 | [0.0,0.0] |
| Urology Service | 0.01 | 0 | 0.16 | [0.0,0.0] |
| | **111** | | | |
| | **Mean** | **Median** | **Std** | **IQR** |
| Cardiology Service | 0.03 | 0 | 0.20 | [0.0,0.0] |
| Cardiothoracic Surgery Service | 0.06 | 0 | 0.33 | [0.0,0.0] |
| Dermatology Service | 0.03 | 0 | 0.21 | [0.0,0.0] |
| Ear Nose and Throat Service | 0.04 | 0 | 0.28 | [0.0,0.0] |
| Elderly Medicine Service | 0.01 | 0 | 0.14 | [0.0,0.0] |
| Gastroenterology Service | 0.03 | 0 | 0.24 | [0.0,0.0] |
| General Internal Medicine Service | 0.03 | 0 | 0.22 | [0.0,0.0] |
| General Surgery Service | 0.03 | 0 | 0.26 | [0.0,0.0] |
| Gynaecology Service | 0.04 | 0 | 0.31 | [0.0,0.0] |
| Neurology Service | 0.03 | 0 | 0.27 | [0.0,0.0] |
| Neurosurgical Service | 0.03 | 0 | 0.21 | [0.0,0.0] |
| Ophthalmology Service | 0.03 | 0 | 0.20 | [0.0,0.0] |
| Oral Surgery Service | 0.05 | 0 | 0.30 | [0.0,0.0] |
| Plastic Surgery Service | 0.03 | 0 | 0.28 | [0.0,0.0] |
| Respiratory Medicine Service | 0.03 | 0 | 0.21 | [0.0,0.0] |
| Rheumatology Service | 0.03 | 0 | 0.26 | [0.0,0.0] |
| Trauma and Orthopaedic Service | 0.03 | 0 | 0.26 | [0.0,0.0] |
| Urology Service | 0.03 | 0 | 0.22 | [0.0,0.0] |
| | **Primary Care Contact** | | | |
| | **Mean** | **Median** | **Std** | **IQR** |
| Cardiology Service | 1.00 | 0 | 2.21 | [0.0,1.0] |
| Cardiothoracic Surgery Service | 1.44 | 0 | 2.69 | [0.0,2.0] |
| Dermatology Service | 0.86 | 0 | 1.96 | [0.0,1.0] |
| Ear Nose and Throat Service | 1.40 | 0 | 2.66 | [0.0,2.0] |
| Elderly Medicine Service | 1.26 | 0 | 2.30 | [0.0,2.0] |

| | | | | |
|---|---|---|---|---|
| Gastroenterology Service | 1.10 | 0 | 2.35 | [0.0,1.0] |
| General Internal Medicine Service | 1.11 | 0 | 2.40 | [0.0,1.0] |
| General Surgery Service | 1.14 | 0 | 2.39 | [0.0,1.0] |
| Gynaecology Service | 1.30 | 0 | 2.56 | [0.0,2.0] |
| Neurology Service | 0.97 | 0 | 2.20 | [0.0,1.0] |
| Neurosurgical Service | 1.14 | 0 | 2.38 | [0.0,1.0] |
| Ophthalmology Service | 1.17 | 0 | 2.38 | [0.0,1.0] |
| Oral Surgery Service | 0.90 | 0 | 2.06 | [0.0,1.0] |
| Plastic Surgery Service | 1.00 | 0 | 2.21 | [0.0,1.0] |
| Respiratory Medicine Service | 1.10 | 0 | 2.33 | [0.0,1.0] |
| Rheumatology Service | 1.07 | 0 | 2.32 | [0.0,1.0] |
| Trauma and Orthopaedic Service | 1.26 | 0 | 2.54 | [0.0,2.0] |
| Urology Service | 1.11 | 0 | 2.39 | [0.0,1.0] |
| | **Prescription** | | | |
| | **Mean** | **Median** | **Std** | **IQR** |
| Cardiology Service | 2.08 | 1 | 5.77 | [0.0,2.0] |
| Cardiothoracic Surgery Service | 2.31 | 1 | 5.89 | [0.0,2.0] |
| Dermatology Service | 1.51 | 0 | 4.40 | [0.0,2.0] |
| Ear Nose and Throat Service | 2.70 | 1 | 7.04 | [0.0,3.0] |
| Elderly Medicine Service | 3.66 | 1 | 7.76 | [0.0,4.0] |
| Gastroenterology Service | 2.26 | 1 | 6.33 | [0.0,2.0] |
| General Internal Medicine Service | 2.30 | 1 | 6.44 | [0.0,2.0] |
| General Surgery Service | 2.27 | 1 | 6.33 | [0.0,2.0] |
| Gynaecology Service | 2.00 | 1 | 5.86 | [0.0,2.0] |
| Neurology Service | 1.67 | 0 | 4.97 | [0.0,2.0] |
| Neurosurgical Service | 2.55 | 1 | 7.12 | [0.0,2.0] |
| Ophthalmology Service | 2.87 | 1 | 7.28 | [0.0,3.0] |
| Oral Surgery Service | 1.22 | 0 | 4.09 | [0.0,1.0] |
| Plastic Surgery Service | 1.90 | 0 | 5.60 | [0.0,2.0] |
| Respiratory Medicine Service | 2.40 | 1 | 6.32 | [0.0,2.0] |
| Rheumatology Service | 2.01 | 1 | 5.72 | [0.0,2.0] |
| Trauma and Orthopaedic Service | 2.80 | 1 | 7.41 | [0.0,3.0] |
| Urology Service | 2.60 | 1 | 6.92 | [0.0,2.0] |
| | **Secondary** | | | |
| | **Mean** | **Median** | **Std** | **IQR** |
| Cardiology Service | 0.30 | 0 | 1.50 | [0.0,0.0] |
| Cardiothoracic Surgery Service | 0.44 | 0 | 2.00 | [0.0,0.0] |
| Dermatology Service | 0.27 | 0 | 1.39 | [0.0,0.0] |
| Ear Nose and Throat Service | 0.43 | 0 | 1.88 | [0.0,0.0] |
| Elderly Medicine Service | 0.34 | 0 | 1.40 | [0.0,0.0] |
| Gastroenterology Service | 0.31 | 0 | 1.53 | [0.0,0.0] |
| General Internal Medicine Service | 0.31 | 0 | 1.50 | [0.0,0.0] |
| General Surgery Service | 0.33 | 0 | 1.62 | [0.0,0.0] |
| Gynaecology Service | 0.45 | 0 | 2.06 | [0.0,0.0] |

| | | | | |
|---|---|---|---|---|
| Neurology Service | 0.30 | 0 | 1.52 | [0.0,0.0] |
| Neurosurgical Service | 0.31 | 0 | 1.48 | [0.0,0.0] |
| Ophthalmology Service | 0.33 | 0 | 1.48 | [0.0,0.0] |
| Oral Surgery Service | 0.34 | 0 | 1.54 | [0.0,0.0] |
| Plastic Surgery Service | 0.29 | 0 | 1.46 | [0.0,0.0] |
| Respiratory Medicine Service | 0.30 | 0 | 1.44 | [0.0,0.0] |
| Rheumatology Service | 0.33 | 0 | 1.63 | [0.0,0.0] |
| Trauma and Orthopaedic Service | 0.35 | 0 | 1.63 | [0.0,0.0] |
| Urology Service | 0.31 | 0 | 1.49 | [0.0,0.0] |

**Table S2** Number of contacts controls had with the health service during the individual study periods.

#### *Is there a difference in health service utilisation?*

| Specialty | 111 | Primary care contact | Primary care prescription | Community | 999 | Secondary | Mental health |
|---|---|---|---|---|---|---|---|
| General Surgery | 0.0 [0.0,0.015] | 0.0 [0.0,0.0] | 0.0 [0.0,0.0] | 0.0 [0.0,0.0] | 0.0 [0.0,0.0] | 0.0 [0.0,0.0] | 0.0 [0.0,0.006] |
| Urology | 0.0 [0.0,0.001] | 0.0 [0.0,0.0] | 0.0 [0.0,0.0] | 0.0 [0.0,0.0] | 0.0 [0.0,0.002] | 0.0 [0.0,0.0] | 0.003 [0.0,0.02] |
| Trauma and Orthopaedic | 0.0 [0.0,0.024] | 0.0 [0.0,0.0] | 0.0 [0.0,0.0] | 0.0 [0.0,0.0] | 0.0 [0.0,0.002] | 0.0 [0.0,0.0] | 0.001 [0.0,0.012] |
| Ear Nose and Throat | 0.0 [0.0,0.006] | 0.0 [0.0,0.0] | 0.0 [0.0,0.0] | 0.104 [0.032,0.309] | 0.0 [0.0,0.001] | 0.0 [0.0,0.0] | 0.001 [0.0,0.041] |
| Ophthalmology | 0.0 [0.0,0.009] | 0.0 [0.0,0.0] | 0.0 [0.0,0.0] | 0.004 [0.0,0.036] | 0.0 [0.0,0.021] | 0.0 [0.0,0.0] | 0.021 [0.001,0.238] |
| Oral Surgery | 0.0 [0.0,0.153] | 0.0 [0.0,0.0] | 0.0 [0.0,0.002] | 0.24 [0.017,0.695] | 0.001 [0.0,0.16] | 0.0 [0.0,0.0] | 0.001 [0.0,0.022] |
| Neurosurgical | 0.011 [0.0,0.36] | 0.0 [0.0,0.0] | 0.0 [0.0,0.0] | 0.001 [0.0,0.006] | 0.0 [0.0,0.007] | 0.0 [0.0,0.0] | 0.042 [0.001,0.37] |
| Plastic Surgery | 0.233 [0.034, 0.783] | 0.0 [0.0,0.0] | 0.014 [0.0,0.22] | 0.063 [0.001,0.581] | 0.349 [0.057,0.807] | 0.0 [0.0,0.0] | 0.25 [0.044,0.69] |
| Cardiothoracic Surgery | 0.361 [0.161, 0.779] | 0.017 [0.003,0.155] | 0.01 [0.003,0.078] | 0.028 [0.022,0.119] | 0.134 [0.091,0.558] | 0.0 [0.0,0.006] | 0.489 [0.161,0.853] |
| General Internal Medicine | 0.023 [0.002, 0.276] | 0.0 [0.0,0.0] | 0.0 [0.0,0.003] | 0.015 [0.012,0.027] | 0.006 [0.001,0.069] | 0.0 [0.0,0.0] | 0.188 [0.033,0.525] |
| Gastroenterology | 0.0 [0.0,0.004] | 0.0 [0.0,0.0] | 0.0 [0.0,0.0] | 0.134 [0.006,0.57] | 0.0 [0.0,0.002] | 0.0 [0.0,0.0] | 0.004 [0.001,0.023] |
| Cardiology | 0.002 [0.0,0.02] | 0.0 [0.0,0.0] | 0.0 [0.0,0.0] | 0.001 [0.0,0.009] | 0.001 [0.0,0.005] | 0.0 [0.0,0.0] | 0.011 [0.001,0.123] |
| Dermatology | 0.057 [0.002, 0.476] | 0.0 [0.0,0.0] | 0.0 [0.0,0.0] | 0.018 [0.003,0.126] | 0.082 [0.005,0.431] | 0.0 [0.0,0.0] | 0.102 [0.008,0.466] |
| Respiratory Medicine | 0.003 [0.0,0.238] | 0.0 [0.0,0.0] | 0.0 [0.0,0.0] | 0.03 [0.0,0.184] | 0.0 [0.0,0.007] | 0.0 [0.0,0.0] | 0.034 [0.002,0.264] |
| Neurology | 0.0 [0.0,0.001] | 0.0 [0.0,0.0] | 0.0 [0.0,0.0] | 0.002 [0.001,0.005] | 0.0 [0.0,0.0] | 0.0 [0.0,0.0] | 0.0 [0.0,0.002] |
| Rheumatology | 0.018 [0.0,0.457] | 0.0 [0.0,0.0] | 0.0 [0.0,0.0] | 0.284 [0.177,0.548] | 0.049 [0.001,0.516] | 0.0 [0.0,0.0] | 0.004 [0.0,0.104] |

| | | | | | | | |
|---|---|---|---|---|---|---|---|
| Elderly Medicine | 0.012 [0.007, 0.113] | 0.656 [0.553,0.762] | 0.318 [0.14,0.613] | 0.018 [0.005,0.535] | 0.009 [0.003,0.108] | 0.002 [0.0,0.055] | 0.072 [0.044,0.404] |
| Gynaecology | 0.0 [0.0,0.063] | 0.0 [0.0,0.0] | 0.0 [0.0,0.0] | 0.0 [0.0,0.017] | 0.0 [0.0,0.056] | 0.0 [0.0,0.0] | 0.008 [0.0,0.126] |

**Table S3** Results of bootstrapped t-tests to assess whether there is a significant difference in weekly activity of treatments and controls more than 18 weeks after a referral is made. Values correspond to median [CI] p-values.

*How large is the difference in health service utilisation?*

| | 111 | |
|---|---|---|
| **Specialty** | **Median** | **IQR** |
| Cardiology Service | 0.0 | [0.0,0.0] |
| Cardiothoracic Surgery Service | 0.0 | [0.0,0.0] |
| Dermatology Service | 0.0 | [0.0,0.0] |
| Ear Nose and Throat Service | 0.0 | [0.0,0.0] |
| Elderly Medicine Service | 0.0 | [0.0,0.0] |
| Gastroenterology Service | 0.0 | [0.0,0.0] |
| General Internal Medicine Service | 0.0 | [0.0,0.0] |
| General Surgery Service | 0.0 | [0.0,0.0] |
| Gynaecology Service | 0.0 | [0.0,0.0] |
| Neurology Service | 0.0 | [0.0,0.0] |
| Neurosurgical Service | 0.0 | [0.0,0.0] |
| Ophthalmology Service | 0.0 | [0.0,0.0] |
| Oral Surgery Service | 0.0 | [0.0,0.0] |
| Plastic Surgery Service | 0.0 | [0.0,0.0] |
| Respiratory Medicine Service | 0.0 | [0.0,0.0] |
| Rheumatology Service | 0.0 | [0.0,0.0] |
| Trauma and Orthopaedic Service | 0.0 | [0.0,0.0] |
| Urology Service | 0.0 | [0.0,0.0] |
| | **Primary Care Contact** | |
| | **Median** | **IQR** |
| Cardiology Service | 0.0 | [0.0,11.8] |
| Cardiothoracic Surgery Service | 5.5 | [-0.3,15.9] |
| Dermatology Service | 0.0 | [0.0,10.3] |
| Ear Nose and Throat Service | 2.0 | [-0.3,10.0] |
| Elderly Medicine Service | 0.0 | [-2.5,8.6] |
| Gastroenterology Service | 0.0 | [0.0,12.0] |
| General Internal Medicine Service | 3.1 | [0.0,15.5] |
| General Surgery Service | 0.0 | [0.0,11.9] |
| Gynaecology Service | 0.0 | [0.0,10.6] |
| Neurology Service | 0.0 | [0.0,12.4] |
| Neurosurgical Service | 2.0 | [0.0,12.6] |
| Ophthalmology Service | 0.0 | [-2.4,8.7] |
| Oral Surgery Service | 0.0 | [0.0,5.1] |
| Plastic Surgery Service | 0.0 | [0.0,9.3] |
| Respiratory Medicine Service | 0.0 | [0.0,14.0] |
| Rheumatology Service | 0.0 | [0.0,12.3] |
| Trauma and Orthopaedic Service | 2.0 | [0.0,11.4] |
| Urology Service | 0.0 | [0.0,12.0] |
| | **Primary Care Prescription** | |
| | **Median** | **IQR** |
| Cardiology Service | 2.0 | [-4.6,26.5] |

| | | |
|---|---|---|
| Cardiothoracic Surgery Service | 17.3 | [-1.1,34.1] |
| Dermatology Service | 0.0 | [-2.4,15.2] |
| Ear Nose and Throat Service | 2.0 | [-3.5,15.6] |
| Elderly Medicine Service | 3.3 | [-9.8,34.5] |
| Gastroenterology Service | 5.7 | [-0.7,28.9] |
| General Internal Medicine Service | 6.9 | [-1.2,30.5] |
| General Surgery Service | 2.0 | [-3.0,22.4] |
| Gynaecology Service | 0.0 | [-2.8,13.8] |
| Neurology Service | 2.7 | [0.0,27.2] |
| Neurosurgical Service | 8.0 | [-0.2,39.1] |
| Ophthalmology Service | 0.0 | [-8.8,23.8] |
| Oral Surgery Service | 0.0 | [-1.2,5.7] |
| Plastic Surgery Service | 0.0 | [-3.1,15.8] |
| Respiratory Medicine Service | 4.7 | [-2.4,33.0] |
| Rheumatology Service | 4.3 | [0.0,27.1] |
| Trauma and Orthopaedic Service | 4.0 | [-2.7,26.7] |
| Urology Service | 4.0 | [-2.3,28.1] |
| | **Community** | |
| | **Median** | **IQR** |
| Cardiology Service | 0.0 | [0.0,0.0] |
| Cardiothoracic Surgery Service | 0.0 | [0.0,0.0] |
| Dermatology Service | 0.0 | [0.0,0.0] |
| Ear Nose and Throat Service | 0.0 | [0.0,0.0] |
| Elderly Medicine Service | 0.0 | [0.0,0.0] |
| Gastroenterology Service | 0.0 | [0.0,0.0] |
| General Internal Medicine Service | 0.0 | [0.0,0.0] |
| General Surgery Service | 0.0 | [0.0,0.0] |
| Gynaecology Service | 0.0 | [0.0,0.0] |
| Neurology Service | 0.0 | [0.0,0.0] |
| Neurosurgical Service | 0.0 | [0.0,0.0] |
| Ophthalmology Service | 0.0 | [0.0,0.0] |
| Oral Surgery Service | 0.0 | [0.0,0.0] |
| Plastic Surgery Service | 0.0 | [0.0,0.0] |
| Respiratory Medicine Service | 0.0 | [0.0,0.0] |
| Rheumatology Service | 0.0 | [0.0,0.0] |
| Trauma and Orthopaedic Service | 0.0 | [0.0,0.0] |
| Urology Service | 0.0 | [0.0,0.0] |
| | **999** | |
| | **Median** | **IQR** |
| Cardiology Service | 0.0 | [0.0,0.0] |
| Cardiothoracic Surgery Service | 0.0 | [0.0,0.0] |
| Dermatology Service | 0.0 | [0.0,0.0] |
| Ear Nose and Throat Service | 0.0 | [0.0,0.0] |
| Elderly Medicine Service | 0.0 | [0.0,0.0] |

| | | |
|---|---|---|
| Gastroenterology Service | 0.0 | [0.0,0.0] |
| General Internal Medicine Service | 0.0 | [0.0,0.0] |
| General Surgery Service | 0.0 | [0.0,0.0] |
| Gynaecology Service | 0.0 | [0.0,0.0] |
| Neurology Service | 0.0 | [0.0,0.0] |
| Neurosurgical Service | 0.0 | [0.0,0.0] |
| Ophthalmology Service | 0.0 | [0.0,0.0] |
| Oral Surgery Service | 0.0 | [0.0,0.0] |
| Plastic Surgery Service | 0.0 | [0.0,0.0] |
| Respiratory Medicine Service | 0.0 | [0.0,0.0] |
| Rheumatology Service | 0.0 | [0.0,0.0] |
| Trauma and Orthopaedic Service | 0.0 | [0.0,0.0] |
| Urology Service | 0.0 | [0.0,0.0] |
| | **Secondary** | |
| | **Median** | **IQR** |
| Cardiology Service | 0.0 | [0.0,7.404] |
| Cardiothoracic Surgery Service | 17.9 | [4.3,33.8] |
| Dermatology Service | 0.0 | [0.0,5.9] |
| Ear Nose and Throat Service | 0.0 | [0.0,6.4] |
| Elderly Medicine Service | 0.0 | [0.0,6.6] |
| Gastroenterology Service | 2.0 | [0.0,11.4] |
| General Internal Medicine Service | 4.0 | [0.0,12.8] |
| General Surgery Service | 2.0 | [0.0,11.1] |
| Gynaecology Service | 0.0 | [0.0,8.7] |
| Neurology Service | 0.0 | [0.0,8.0] |
| Neurosurgical Service | 4.0 | [0.0,12.1] |
| Ophthalmology Service | 0.0 | [0.0,8.0] |
| Oral Surgery Service | 0.0 | [0.0,0.0] |
| Plastic Surgery Service | 2.3 | [0.0,11.9] |
| Respiratory Medicine Service | 5.9 | [0.0,16.0] |
| Rheumatology Service | 0.0 | [0.0,8.4] |
| Trauma and Orthopaedic Service | 4.0 | [0.0,11.8] |
| Urology Service | 2.9 | [0.0,11.9] |
| | **Mental Health** | |
| | **Median** | **IQR** |
| Cardiology Service | 0.0 | [0.0,0.0] |
| Cardiothoracic Surgery Service | 0.0 | [0.0,0.0] |
| Dermatology Service | 0.0 | [0.0,0.0] |
| Ear Nose and Throat Service | 0.0 | [0.0,0.0] |
| Elderly Medicine Service | 0.0 | [0.0,0.0] |
| Gastroenterology Service | 0.0 | [0.0,0.0] |
| General Internal Medicine Service | 0.0 | [0.0,0.0] |
| General Surgery Service | 0.0 | [0.0,0.0] |
| Gynaecology Service | 0.0 | [0.0,0.0] |

| | | |
|---|---|---|
| Neurology Service | 0.0 | [0.0,0.0] |
| Neurosurgical Service | 0.0 | [0.0,0.0] |
| Ophthalmology Service | 0.0 | [0.0,0.0] |
| Oral Surgery Service | 0.0 | [0.0,0.0] |
| Plastic Surgery Service | 0.0 | [0.0,0.0] |
| Respiratory Medicine Service | 0.0 | [0.0,0.0] |
| Rheumatology Service | 0.0 | [0.0,0.0] |
| Trauma and Orthopaedic Service | 0.0 | [0.0,0.0] |
| Urology Service | 0.0 | [0.0,0.0] |

**Table S4** The amount of additional health service utilisation of patients waiting for treatment.

Values represent the number of additional contacts per year.
